# MULTI-OMICS TEMPORAL PROFILING OF AXIAL SPONDYLOARTHRITIS PATIENTS REVEALS AN ASSOCIATION OF THERAPEUTIC RESPONSE TO ADALIMUMAB WITH DISEASE ACTIVITY AND INNATE / ADAPTIVE IMMUNITY

**DOI:** 10.1101/2022.08.02.22278314

**Authors:** Daniel Sobral, Ana F Fernandes, Atlas Sardoo, Miguel Bernardes, Patrícia Pinto, Helena Santos, João Lagoas-Gomes, José Tavares-Costa, José AP Silva, João M Dias, Alexandra Bernardo, Jean-Charles Gaillard, Jean Armengaud, Vladimir Benes, Lúcia Domingues, Nélia Gouveia, Sara Maia, Jaime C Branco, Ana V Coelho, Fernando M Pimentel-Santos

## Abstract

**Background:** Axial Spondyloarthritis can lead to significant disability and impairment in quality of life. TNF inhibitors are recommended to patients enduring active disease despite conventional treatment. Nonetheless, up to 40% of patients of patients fail to respond to TNF inhibitors. In this context, it is important to identify as early as possible patients highly likely to respond. This study aims at identifying, among axial spondyloarthritis patients undergoing treatment with the TNF inhibitor adalimumab, early molecular biomarkers differentiating good responders from non-responders after 14 weeks of treatment, as measured by ASAS20.

**Methods:** Peripheral blood RNA sequencing and serum proteins measured by mass spectrometry were evaluated in a cohort of biologic naïve axial spondyloarthritis patients (n = 35), before (baseline) and after (3-5 days, 2 weeks and 14 weeks) treatment with adalimumab. Results from differential expression analysis were used in combination with clinical data to build logistic regression models and random forest models to predict response to adalimumab at baseline.

**Results:** Responders to adalimumab presented higher levels of markers of innate immunity at baseline, mostly related with neutrophils, and lower levels of adaptive immunity markers, particularly B-cells. A logistic regression model incorporating ASDAS-CRP and AFF3, the top differentially expressed gene between responders and non-responders at baseline, enabled an accurate prediction of response to adalimumab in our cohort (AUC=0.96), with random forest models suggesting 80% predictive accuracy. A treatment-associated signature suggests a reduction in inflammatory activity, with C-reactive protein and Haptoglobin showing strong and early decrease in the serum of axial spondyloarthritis patients, while a cluster of apolipoproteins showed increased expression at week 14.

**Conclusions:** Differences in disease activity and/or blood innate/adaptive immune cell type composition at baseline may be a major contributor to response to adalimumab in axial spondyloarthritis, where a model including clinical and blood gene expression variables shows high predictive power. Our results suggest novel molecular biomarkers of response to adalimumab at baseline.

**Trial registration:** Axial spondyloarthritis patients were selected from participants of the Bioefficacy study - Biomarkers Identification of Anti-TNFα Agent’s Efficacy in Ankylosing Spondylitis Patients Using a Transcriptome Analysis and Mass Spectrometry (clinical trials.gov identifier NCT02492217).

## BACKGROUND

Axial Spondyloarthritis (axSpA) can lead to significant disability and impairment in quality of life [1]. Inflammatory back pain is a characteristic symptom, and new bone formation with syndesmophytes and ankylosis are the disease radiographic hallmarks. Clinical features of axSpA are heterogeneous, including inflammatory back pain, asymmetrical peripheral oligoarthritis (predominantly of the lower limbs), enthesitis, and specific organ involvement such as acute anterior uveitis, psoriasis, and chronic inflammatory bowel disease. Pulmonary, renal, neurological, aortic root involvement and conduction abnormalities are all rare complications of axSpA [2].

In axSpA, non-steroidal anti-inflammatory drugs (NSAIDs) have a central role in treatment and are considered the first-choice drug treatment. However, biological disease-modifying antirheumatic drugs (bDMARDs) including TNF inhibitors (TNFi), are recommended to patients enduring active disease despite conventional treatment (or intolerance/contraindication) [3]. The efficacy of TNFi has been documented in several studies showing significant and early improvements in disease activity and function [4] sustainable for long periods of time [5]. In spite of its well documented benefit in axSpA, up to an estimated 40% axSpA patients fail to respond to TNFi treatment [6] or complain with adverse events [7].

The concept of “window of opportunity” is of critical importance in rheumatoid arthritis (RA) [8] and seems to be also relevant in the axSpa context, with studies of magnetic resonance imaging in the context of TNFi treatment suggesting that early effective suppression of inflammation has the potential to reduce radiographic damage [9]. Patients that fail to respond to the first bDMARD usually switch to another bDMARD (with the same or other mechanism of action, such as IL-17 inhibitors [10]), and it may take several iterations to find a drug that reduces disease activity [3]. Response to an effective therapy can take several weeks/months, and the delay caused by non-response may imply irreversible damage. In this context, it is important to identify as early as possible the patients highly likely to respond, meaning to achieve remission or low disease activity, following the treat to target concept [11].

Studies specifically in axSpA indicate that primary non-responders to TNFi tend to be older, HLA-B27 negative, with longer disease duration (> 20 years), higher structural damage and poor function [12]. Likewise, good response to therapy was associated with younger age, HLA-B27 carriage, short disease duration (<10 years), elevation of acute phase reactants (CRP), and marked spinal inflammation (detectable by MRI) [13]. Higher levels of CRP is the most common marker associated with good response, but higher levels of other inflammation markers such as IL6 [14] and calprotectin [15] have also been associated with positive outcomes of TNFi treatment.

The molecular characterization of anti-TNF response in axSpA revealed the unsurprising involvement of genes related with immunity and inflammation [16,17]. Although several studies address the overall effect of TNFi treatment in the axSpA context [18], very few specifically address the molecular changes associated with response/non-response to anti-TNF treatment in axSpA, most of them focusing on specific markers of inflammation in the sera of a limited set of patients. In particular, no study systematically assessed large scale transcriptomics and/or proteomics data to find early predictors of response to anti-TNF in axSpA. On the other hand, several studies have tried to develop such predictors in RA, with variable success [19–21]. One study using whole blood transcriptomics achieved 65% accuracy of response to infliximab in RA with a 10-gene biomarker set [19]. In another study using transcriptomics of monocytes, CD11c was found to be a very good predictor of response (95% accuracy) to adalimumab monotherapy in RA [20]. Thomson and colleagues used publicly available data to develop a model including 18 signaling mechanisms that could increase the capacity to discover non-responders, from 27% to 59% [21]. More recently, several studies suggested an interplay between innate and adaptive immunity, with a higher myeloid-driven inflammation in responders and higher lymphoid activity in non-responders [21–23].

Reliable predictors of outcome for anti-TNF monotherapies in axSpA are currently not yet available. The goal of this study is thus the identification of baseline predictors, using transcriptomic and proteomic approaches, of patient response to anti-TNF therapy (adalimumab) in axSpA using peripheral blood samples, which are particularly appealing given the relative ease of obtaining samples as part of patient follow-up.

## METHODS

### Study design and samples collection

AxSpa patients were selected from participants of the Bioefficacy study - Biomarkers Identification of Anti-TNFα Agent’s Efficacy in Ankylosing Spondylitis Patients Using a Transcriptome Analysis and Mass Spectrometry (clinical trials.gov identifier NCT02492217). This is a multicentric, prospective, nonrandomized, 14-weeks study that includes axSpA adult patients according to axSpA ASAS criteria [24]. The study included biologic naïve patients, starting TNFi therapy with adalimumab (40mg subcutaneously fortnightly), according to the Portuguese Society of Rheumatology Guidelines [25] (see supplementary material). Clinical evaluations and peripheral blood collections were performed at baseline (start bDMARD), and after 3-5 days, 2 weeks and 14 weeks. Patients were classified as responders and non-responders, according to ASAS20 [26,27] at week 14. To have 80% power to detect a 0.5SD difference between groups at p=0.05 (paired t-test), we estimated that we would need samples from 18 responders and 18 non-responders. Usually only 60% of patients after starting TNFi reach ASAS20, which means that we would need to include a larger number of patients to establish the subgroups for analysis. Thus, we included the number of patients necessary to ensure 18 non-responders, after which we closed the recruitment period. All clinical evaluations were performed by previously trained rheumatologists. Blood samples were collected from all subjects at baseline to test for HLA-B27 status and at each timepoint to determine C-reactive protein (CRP), Erythrocyte Sedimentation Rate (ESR), other biochemical parameters and for RNA-seq and serum proteomic analysis.

### RNA preparation and NGS sequencing (RNA-seq)

Peripheral blood samples were collected into PAXgene Blood RNA System® tubes and stored at -80° C according to the manufacturer’s recommendations [28]. Total RNA was extracted from whole blood samples according to the standard PAXgene protocol (Qiagen, 2008). The quantity of RNA was measured using a NanoDrop 2000/2000c Spectrophotometer according to the manufacturer’s procedure (Thermo-Scientific, 2000); RNAs with a 260:280 ratio of ≥1.5 were sequenced as below. The quality and quantity of the libraries was assessed by Fragment Analyzer with the method of DNF-474-22 - HS NGS Fragment 1-6000bp (Agilent). Sequencing library preparation was performed using Illumina TruSeq stranded mRNA library preparation kits, with 100ng of total RNA as input. Libraries were sequenced on an Illumina NextSeq500 sequencer (average of 39 million reads per sample, 75 base-pair paired-end). Sample correspondence between timepoints was confirmed using SmaSH [29]. We also used the transcriptomic data to confirm gender and HLA-B27 status.

### RNA-Seq data analysis

Raw sequencing reads were aligned to gencode (v32) transcripts using kallisto (version 0.46.1) [43], reaching an average of 86% reads assigned to genes (gene counts are in Supp. Table 11). The edgeR R package was used to filter low-expressed genes with the filterByExpr function and to normalize raw counts with the trimmed mean of M-values (TMM) normalization approach [44]. The limma R package was used to apply a voom transformation for variance stabilization [45], and to obtain differentially expressed genes through an empirical bayes approach. Genes were considered differentially expressed if the adjusted p-value of the test was less than 0.05. Functional enrichment analysis was performed using the fgsea R package, based on ranks of the moderated t-statistic from the empirical bayes analysis. The per-gene variance explained by each variable was estimated using the variancePartition R package. Permutational multivariate analysis of variance (adonis) was performed using the vegan R package.

### Inference of Immune cell populations from RNA-Seq data

The hemograms collected in each timepoint were used to correlate with data obtained from transcriptomic analysis. As common hemograms did not provide specific information on B-cells and other more specific cell types, we used Cibersort [30] to infer relative frequencies of immune cell populations by comparing normalized log2(counts per million) of the blood transcriptomes to the Abbas et al. signatures [31]. To assess the accuracy of these measurements, we correlated the relative frequencies obtained with RNA-Seq with the values we obtained with clinical hemograms (Supplemental Figure 8A,B, pearson R=0.84 and p=3.3e-8 for Neutrophils, R=0.86 and p=1.2e-8 for Lymphocytes). For consistency, we used only frequencies from Cibersort estimated values in the analyses for Fig4C and Fig2D, and in logistic regression models. In Supp. Fig. 8C,D we used exclusively hemogram data, and in Supp. Fig. 8E.F we used frequencies estimated with Cibersort confirming the same conclusions.

Quantitative set analysis for gene expression (QUSAGE) [32] was used to assess the fold change of immune signature gene sets from Lewis et al. [33] (Supp. Fig. 7A).

### Proteomics Analysis (LC-MS/MS)

#### Immunoaffinity depletion of high-abundance proteins

Peripheral blood samples were collected into Clot Activator Tubes (Monovette Serum Gel Z-7.5 mL, Sarsted) containing 100 μL of Protease Inhibitor Cocktail (Sigma-Aldrich). The six most abundant proteins in serum were depleted using the Multiple Affinity Removal Spin Cartridge Human 6 Kit (Agilent Technologies®) following manufacturer’s instructions. The remaining proteins were concentrated using 4 mL Spin Concentrators with 5000 MWCO (Agilent Technologies®). A centrifugation was performed (with a fixed angle rotor) at 4000 x g and 10ºC until the sample reached a volume between 100 and 140 μL, after which it was recovered from the bottom of the concentrator pocket and stored at -20ºC until further analysis. In order to quantify the amount of protein in each sample, the QuantiPro™ BCA Kit (Sigma-Aldrich®) was used.

#### In-gel protein digestion

50 µg of total proteins was diluted with MilliQ water to a final volume of 20 µL and 10 µL of LDS3X (Invitrogen™ by Life Technologies™) was added, for a final volume of 30 µL. Samples were heated for 5 min at 99ºC and briefly centrifuged (16,000 g for 1 min). The whole volume of the supernatant containing the soluble proteins was loaded on a NuPAGE 4-12% Bis-Tris (Invitrogen™ by Life Technologies™) gel and the proteins were subjected to SDS-PAGE electrophoresis for 5 min. After migration, the gels were stained with Coomassie SimplyBlue SafeStain (Invitrogen™ by Life Technologies™) for 5 min and washed with water overnight with gentle agitation. Polyacrylamide bands containing the stained proteome were cut by the limit of gel wells, between the front of migration and the well bottom. Each sample was treated and proteolyzed with trypsin Gold (Promega©) in presence of ProteaseMax detergent (Promega©) as previously described [46]. The final volume of peptide extract was 50 µL.

#### LC-MS/MS analysis

Tryptic peptides were analyzed with a Q-Exactive™ HF high resolution tandem mass spectrometer (ThermoFisher Scientific™) incorporating an ultra-high-field Orbitrap analyser as previously described [34]. Shortly, 10 μL of the resulting peptide mixtures for each sample were injected in a random order. First, peptides were desalted online on a reverse phase precolumn Acclaim PepMap 100 C18 (5 μm, 100 Å, 300 μm id x 5 mm), and then, they were resolved on a reverse phase column Acclaim PepMap 100 C18 (3 μm, 100 Å, 75 μm id x 500 mm) at a flow rate of 200 nL/min with a 90 min gradient of 4 to 25 % of B in 75 min and 25 to 40% of B in 15 min (being A: 0.1% HCOOH and B: 80% CH3CN, 0.1% HCOOH). The Q-Exactive HF instrument was operated according to a Top20 data-dependent method consisting in a scan cycle initiated with a full scan of peptide ions in the ultra-high-field Orbitrap analyzer, followed by serial selection of each of the 20 most abundant precursor ions, high energy collisional dissociation and MS/MS scans. Full scan mass spectra were acquired from m/z 350 to 1,500 with a resolution of 60,000. A peptide exclusion list was established for the most abundant immunodepleted proteins: serum albumin (https://www.uniprot.org/uniprot/P02768) complement C3 (P01024), alpha-2-macroglobulin (P01023), and apolipoprotein B-100 (P04114), in order to focus the analysis on the other proteins. Each MS/MS scan was acquired with a threshold intensity of 83.000, on potential charge states of 2+ and 3+ after ion selection performed with a dynamic exclusion of 10 sec, maximum Inclusion Time (IT) of 60 ms and an m/z isolation window of 2.0. MS/MS spectra at a resolution of 15.000 were searched using MASCOT 2.5.1 software (Matrix Science) against the Swissprot Human database downloaded in July 2019 (20.432 Homo sapiens protein sequences). The following parameters were used for MS/MS spectra assignation: full trypsin specificity, maximum of two missed cleavages, mass tolerances of 5 ppm on the parent ion and 0.02 Da on the secondary ions, fixed modification of carbamidomethyl cysteine (+57.0215), and oxidized methionine (+15.9949) and deamidated (NQ) (0.9840) as dynamic modifications.

#### Protein identification and relative quantification

After LC-MS/MS, a bioinformatic analysis was performed where all peptide matches with a MASCOT peptide score below a query identity threshold p-value of 0.05 were filtered and assigned to proteins. A total of 5.453.298 MS/MS spectra were recorded and 1.632.427 spectra were assigned to peptide sequences from the protein database – these peptide spectral matches are listed in Supp. Table 12. A protein identification was considered valid when at least two different non-ambiguous peptides were detected in the whole dataset. False discovery rate (FDR) for proteins was below 1% when applying these rules with the MASCOT decoy search mode. A total of 333 polypeptide sequences were identified based on at least 2 non-ambiguous peptides – from the initially 377 polypeptide sequences identified, 44 contaminant proteins (keratin and keratin associated proteins) were excluded from further analysis. For each validated protein (listed in Supp. Table 12), the number of MS/MS spectra for all detected non-ambiguous peptides or ‘Spectral Count’ (SC) [47] was used as a proxy of their abundances [48]. To further assess the value of SC as a measure of protein abundance, we compared clinically determined CRP levels with CRP levels measured by proteomics, and found these to be highly correlated (Supp. Fig. 9, pearson R=0.73, spearman rho=0.79, p<2e-16). Differential protein analysis was performed similarly to the transcriptomics, using the SC values as counts.

### Data analysis

Descriptive statistics were used to summarize baseline characteristics for responders and non-responders. Two sample Wilcoxon tests (continuous variables) and chi-square tests of association (categorical variables) were used to compare characteristics between responders and non-responders at different timepoints, in particular between baseline and week 14.

Differential gene and protein expression analysis used the limma R package to apply a voom transformation for variance stabilization on normalized expression values, and to obtain differentially expressed genes through an empirical bayes approach, followed by multiple test correction with the Benjamini-Hochberg method. Genes were considered differentially expressed if the adjusted *p*-value of the test was less than 0.05. Logistic regression models, and plotting was performed using the R software. Sparse partial least squares discriminant analysis (sPLS-DA) was performed using the mixOmics R package. Random forest models were obtained using the randomForest R package.

## RESULTS

### The TNF inhibitor adalimumab induced a reduction in disease activity of most axSpa patients

According to the ASAS20 criteria we obtained 18 non-responders, and selected, from the whole cohort, 18 responders matched to non-responders by age and gender (one non-responder was later excluded as it had missing information). Table 1 briefly summarizes the clinical characteristics of this sub-group of 35 patients included in this study. Responders presented higher values of C-reactive protein (CRP) (p=0.011) and ASDAS-CRP (p<0.001) at baseline (BL). Responders had a higher proportion of HLA-B27 (p=0.01), with 83% having the allele against only 41% of non-responders. Disease activity decreased from baseline to week 14 (w14) in both responders (ASDAS-CRP: from 4.2 at baseline to 1.6 at week 14, p=2e-04; BASDAI: from 6.5 to 1.9, p=2e-04) and non-responders (ASDAS-CRP: from 3.2 to 2.5, p=5e-04; BASDAI: from 5.3 to 4.0, p=6e-03) (Table 1, Supp. Fig. 1A,B). This suggests that treatment with TNFi, with a few exceptions, has lowered inflammatory markers and disease activity scores in most patients.

**Table 1.**
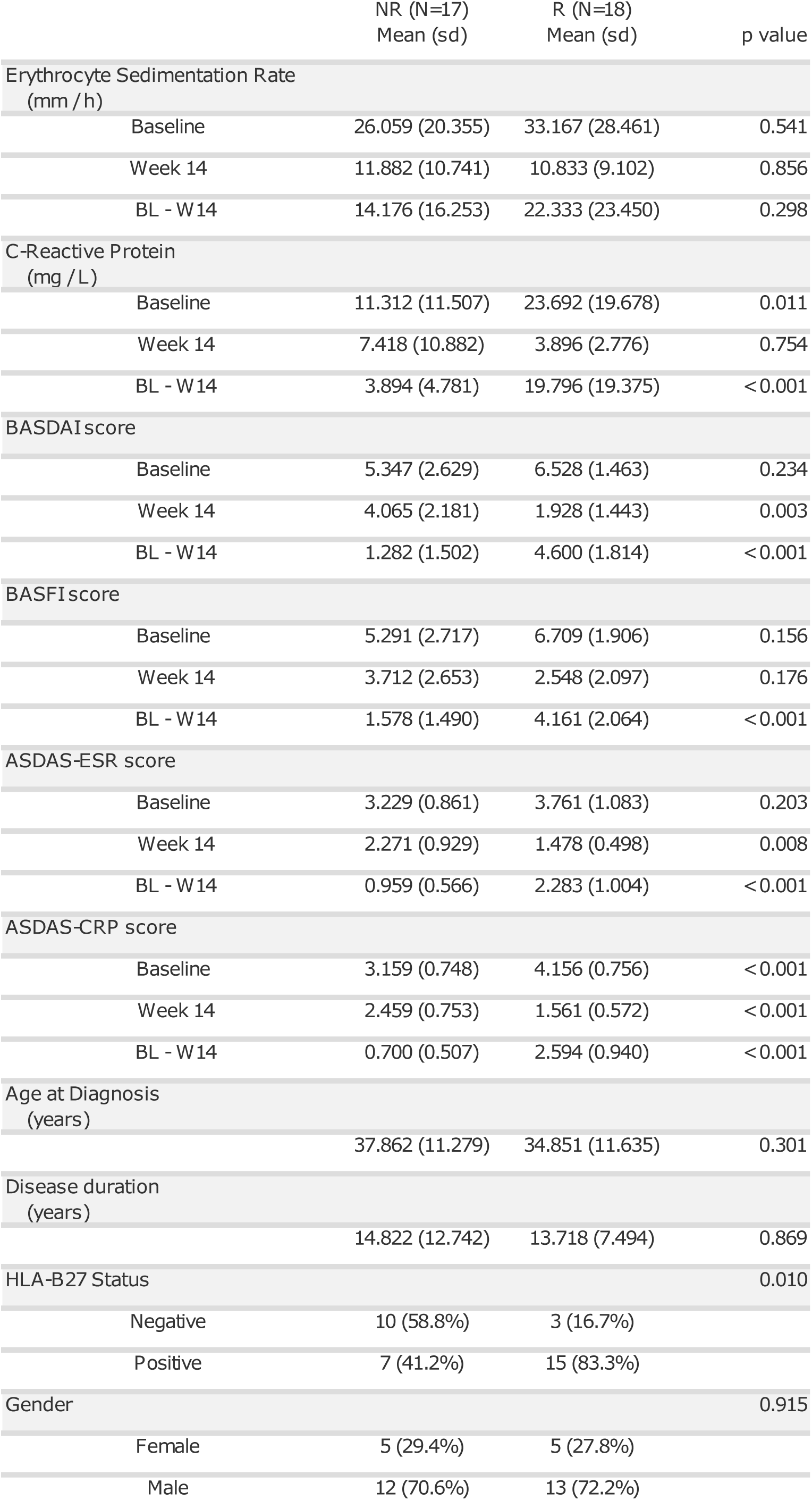
Summary of the clinical characteristics of the cohort. For each continuous variable, the mean and standard deviation within each group were calculated. Two sample Wilcoxon tests (continuous variables) and chi-square tests of association (categorical variables) were used to compare characteristics between “Responders” and “Non-Responders”. Variables include Erythrocyte Sedimentation Rate (ESR, in mm / h), C-Reactive Protein (CRP, in mg / L), Bath Ankylosing Spondylitis Disease Activity Index (BASDAI) scores, Bath Ankylosing Spondylitis Functional Index (BASFI) scores, Ankylosing Spondylitis Disease Activity Score (ASDAS) using the ESR levels (ASDAS-ESR) or CRP levels (ASDAS-CRP). For these characteristics, the value at baseline and week 14 is provided, as well as the difference between the two endpoints. Other fixed clinical characteristics include age at diagnosis (in years of age), disease duration (in years since start of first symptoms), presence (Positive) or absence (Negative) of the HLA-B27 allele, and sex (biological gender) – Female or Male.

### Treatment with adalimumab had a significant impact on the expression of blood cell transcripts and serum proteins of axSpa patients

Expression levels of blood cell transcripts and abundances of serum proteins in axSpA patients did not clearly separate responders from non-responders in an unsupervised principal component analysis (PCA) at neither BL nor at w14 (Fig. 1A). Nonetheless, serum proteomics showed clear differences between BL and w14 in responders, suggesting an effective impact of adalimumab treatment. Indeed, a sparse partial least squares discriminant analysis (sPLS-DA) supports a separation between BL and W14, for both responders and non-responders, not only in proteomics (Supp. Fig. 2), but also in transcriptomics (p<0.05, Fig. 1B). Permutational multivariate analysis of variance indicates that both time points (3% and 17%) and response groups (2% and 4%) can explain a small but statistically significant (p<0.05) part of the observed global variation both in transcript and protein levels, respectively. Moreover, sPLS-DA analysis supports a separation between responders and non-responders at baseline (p<0.01, Fig. 1C). This suggests that treatment with TNFi had a significant impact in the expression of blood cell transcripts and serum proteins of axSpA patients undergoing treatment with adalimumab. Moreover, it also suggests the existence of detectable differences between responders and non-responders at baseline.

**Figure 1:**
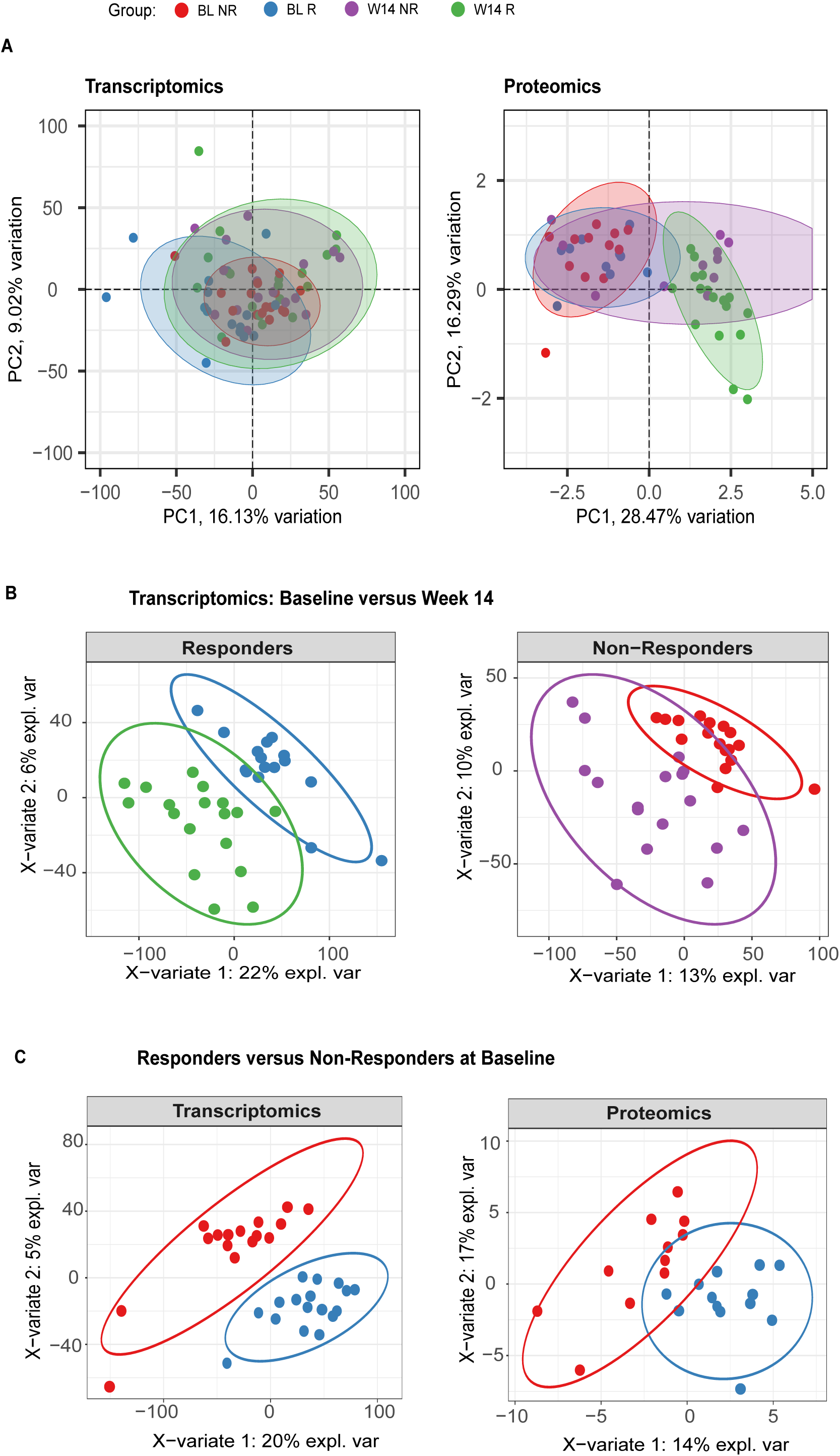
Response to TNFi has a significant impact on the relative abundance of blood cells transcripts and serum proteins of patients. **A**) Principal Component Analysis (PCA) of the blood cell transcriptomics and proteomics data for responders (R) and non-responders (NL) at baseline (BL) and week 14 (W14). For visual clarity two outliers are out of view in the transcriptomics PCA, but all data was used to generate the plot; **B)** Sparse partial least squares discriminant analysis (sPLS-DA) of transcriptomics data, using time as a variable of interest, in responders (AUC=0.99, permutation test p=8e-07) and non-responders (AUC=1, p=6.4e-07); **C)** sPLS-DA of transcriptomics (AUC=1, p=9.6e-07) and proteomics (AUC=1, p=3.4e-05) data at baseline, using response group as a variable of interest. In all cases, AUC and p-value correspond to the two best components of the sPLS-DA. In all graphs, ellipses represent 95% confidence intervals.

### Blood transcriptome data at baseline suggests that response to adalimumab derived from an interplay between innate and adaptive immunity

We tested for differences in gene expression or protein abundance between responders and non-responders at baseline. No proteins (of 112) were found to be significantly differentially abundant between the two groups at BL (Supp. Table 9), but we could detect 92 (of 18688) genes (12 with FC>2) differentially expressed between responders and non-responders at BL, with 16 (0 with FC>2) more expressed in responders and 76 (12 with FC>2) more expressed in non-responders (Fig. 2A, Supp. Table 10). Genes more expressed in responders are associated with inflammation, such as neutrophil degranulation and interferon signaling, while genes more expressed in non-responders are associated with lymphocyte activation, namely B-cell activity, and metabolism, namely translation (Fig. 2B). The top differentially expressed genes are *PAX5, MS4A1* (CD20), *FCRLA, BANK1* and *AFF3*, all associated with B-cells and all significantly more expressed in non-responders at BL (Fig. 2C). Corroborating this observation, estimation of lymphocyte population frequencies using RNA-Seq indicates significantly higher frequencies of B-cells in non-responders at BL (Fig. 2D). Genes associated with B-cells show the strongest overall difference between responders and non-responders at BL. Moreover, there is a significant positive correlation between disease activity and estimated neutrophil frequencies, and a negative correlation between disease activity and estimated B- and T-cell frequencies at BL (Supp. Fig. 7B). Thus, our results suggest that response to adalimumab derives from alterations in the balance between innate and adaptive immunity, indicating an opposing role particularly between neutrophils and B-cells.

**Figure 2:**
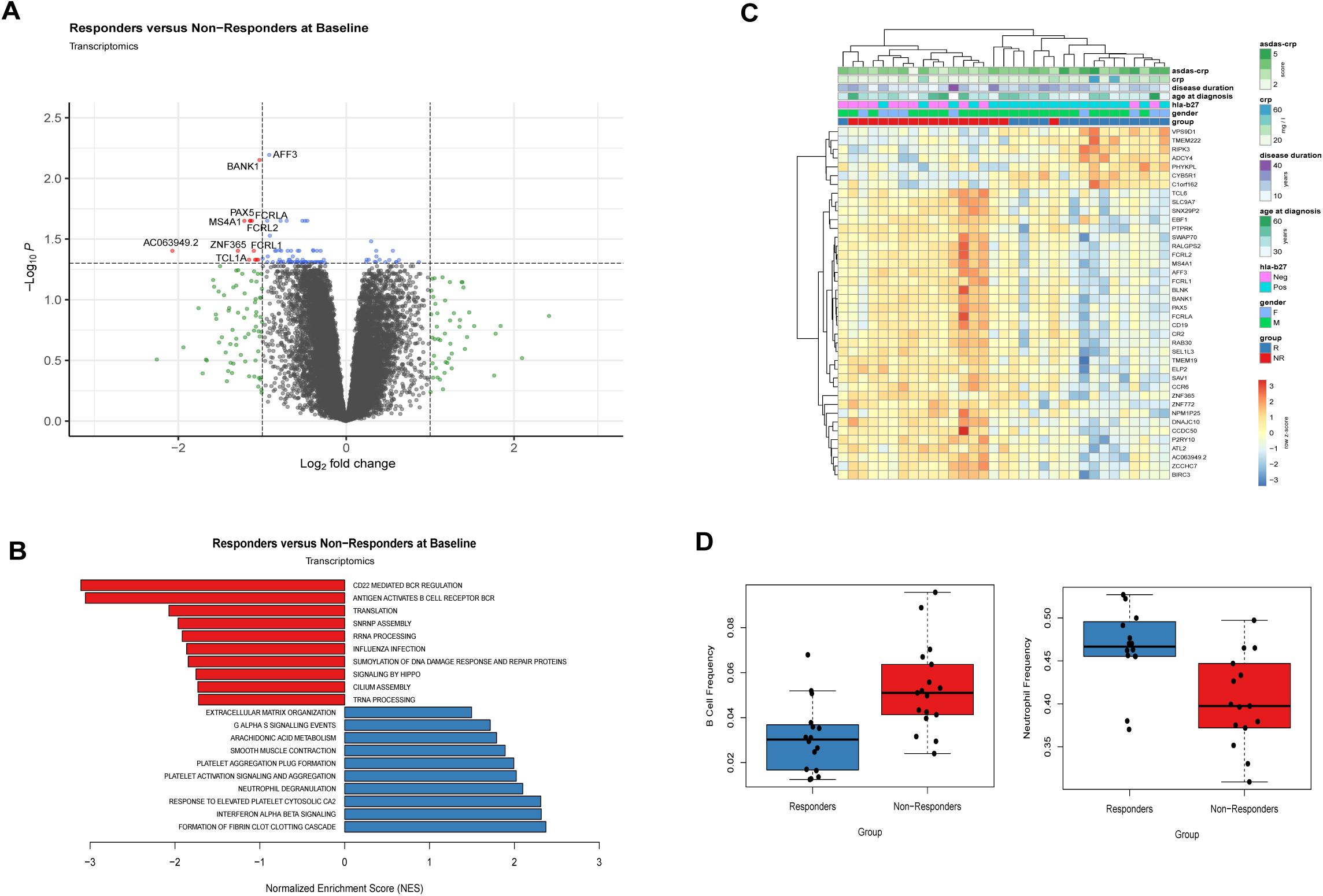
Blood transcriptome data at baseline suggests response to adalimumab derives from an interplay between innate and adaptive immunity. **A)** Volcano plot (log2 of the fold change versus -log10 of the false discovery rate (FDR)) comparing the transcriptomics responder versus non-responder samples at baseline (Supp. Table 10); **B)** Barplot displaying the Normalized Enrichment Score (NES) of representative significant pathways resulting from a gene set enrichment analysis (GSEA) comparing the gene expression of responder versus non-responder samples at baseline; **C)** Heatmap representation of the expression profile of the top 40 differentially expressed genes comparing responder versus non-responder samples at baseline; for visualization purposes, expression values of each gene were scaled towards a standard distribution (z-score); rows and columns were clustered by correlation; **D)** Estimated B-cell and Neutrophil frequencies in responder and non-responder samples at baseline;

### Blood transcriptome data improved ability to differentiate responders versus non-responders at baseline

In our cohort, ASDAS-CRP at baseline was significantly (p=0.011) associated with response in a multivariate logistic model of association with TNFi response (Fig. 3A). HLA-B27 status was also significant (p=0.034), while age at diagnosis and disease duration did not reach statistical significance. ASDAS-CRP was more elevated in responders, with an optimal threshold of 4.15 (100% sensitivity and 50% specificity) when considered in isolation (area under the curve (AUC) = 0.83, Fig. 5B). A model incorporating simultaneously the clinical parameters ASDAS-CRP and HLA-B27 reached an AUC of 0.88. Interestingly, a model replacing HLA-B27 with the ratio between neutrophils and total lymphocytes (N/L) achieved an AUC of 0.84, hinting at the potential for the use of hemogram data as a simple alternative to predict response to treatment. Finally, we incorporated in our models variables from the transcriptomic data. For this, we chose the most robustly differentially expressed gene between responders and non-responders at baseline, which was AFF3, a tissue-restricted nuclear transcriptional activator preferentially expressed in lymphoid tissues. This gene presented the lowest false discovery rate (0.006), and a fold change of 1.9 (higher in non-responders, Fig. 2A,B). Adding the gene expression values of *AFF3* to a logistic regression model including ASDAS-CRP increased the AUC to 0.96 (Fig. 3B). If we choose the second most robust differentially expressed gene (*BANK1*), we reach a similar AUC of 0.94 (not shown). Moreover, a random forest model using ASDAS-CRP, and *AFF3* achieved a predictive accuracy of 80%, better than using ASDAS-CRP alone (60%) or only clinical variables ASDAS-CRP and HLA-B27 (70%). Thus, our results suggest that blood transcriptome data can improve our ability to differentiate responders from non-responders at baseline, and that simple hemogram data can have valuable clinical application.

**Figure 3:**
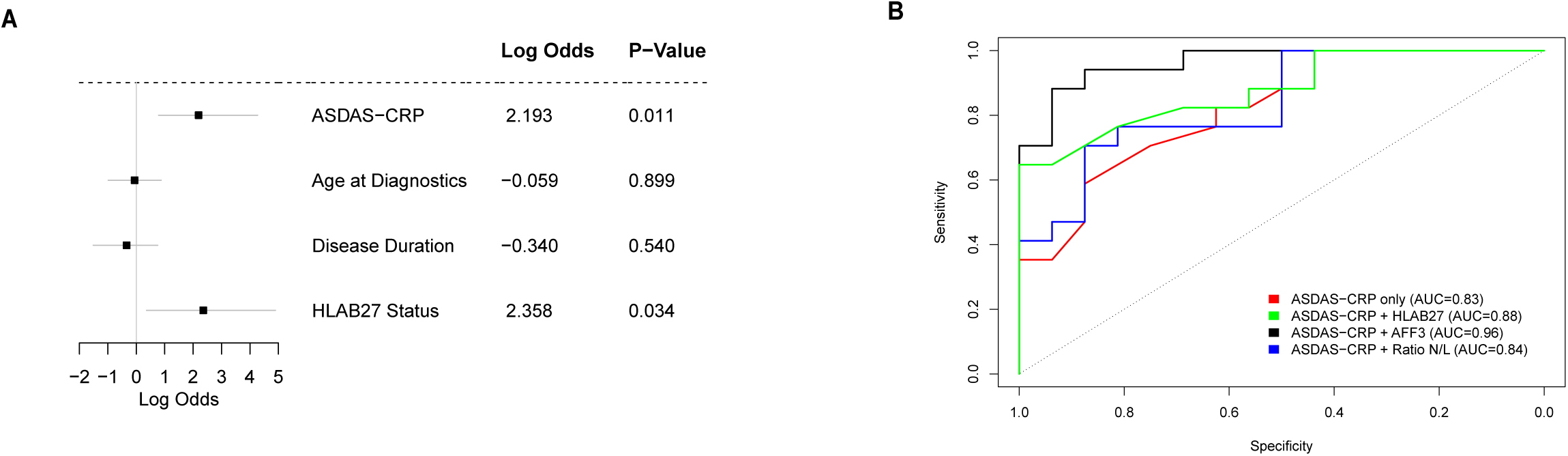
Blood transcriptome data improves ability to differentiate responders versus non-responders at baseline. **A)** Forest plot displaying the logarithm of the odds, 95% confidence interval and p-value of response to adalimumab for different variables from a logistic regression model. **B)** Receiver operating characteristic (ROC) curve displaying specificity and sensitivity for different logistic regression models incorporating: only ASDAS-CRP scores (Area Under the Curve - AUC=0.83); ASDAS-CRP scores and HLA-B27 status (AUC=0.88); ASDAS-CRP scores and the ratio between the estimated frequency of neutrophils and total lymphocytes (N/L, AUC=0.84); ASDAS-CRP and the normalized expression value of *AFF3* (AUC=0.96).

### Transcripts and proteins varying between baseline and week 14 were associated with a decrease in innate immune activity

To assess more concretely the impact of adalimumab treatment, we first looked for differences in gene expression and protein abundance between BL and w14. In responders, 2120 (of 21438) genes (103 genes with fold change (FC) greater than 2) and 41 (of 129) proteins (7 with FC>2) were differentially abundant between BL and w14, of which 1096 genes (41 with FC>2) and 25 proteins (4 with FC>2) were upregulated at w14 (Fig. 4A, Supp. Tables 1,2).

**Figure 4:**
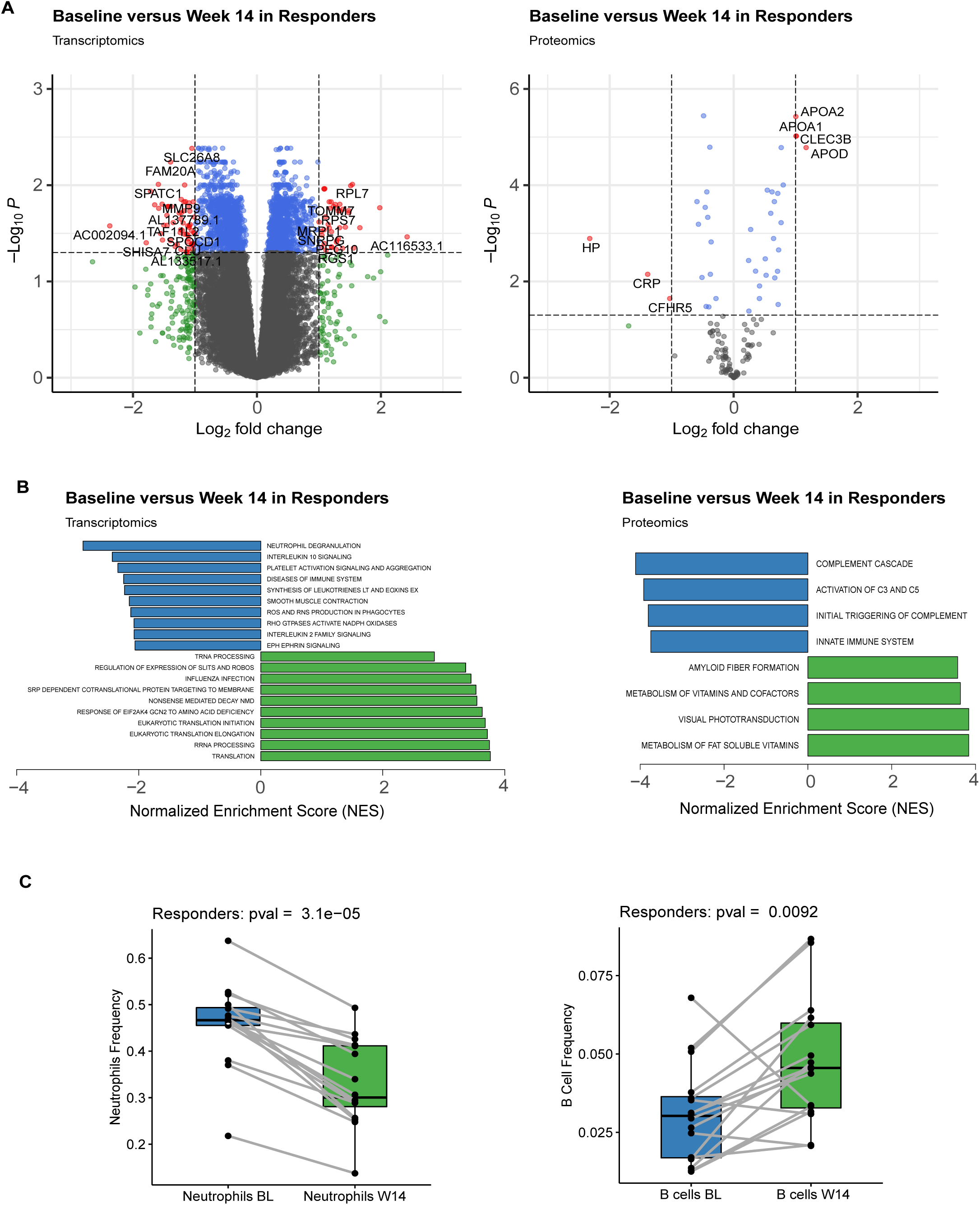
Transcripts and proteins varying between baseline and week 14 were associated with a decrease in innate immune activity. **A)** Volcano plot (log2 of the fold change versus -log10 of the false discovery rate (FDR)) comparing the blood cell transcriptomics (Supp. Table 1) and serum proteomics (Supp. Table 2) baseline samples versus week 14 samples in responders; non-significant (NS) genes/proteins are in grey; in green genes/proteins that are not statistically significant (FDR>0.05) but have an estimated fold change greater than 2; in blue genes/proteins that are statistically significant but have a milder fold change (less than 2); in red genes/proteins that are statistically significant and have a fold change greater than 2. All red proteins and some of the red gene names are displayed in the plot. **B)** Barplot displaying the Normalized Enrichment Score (NES) of representative significant pathways resulting from a gene set enrichment analysis (GSEA) comparing the gene expression or protein abundances of w14 (green) against BL (blue) responder samples; **C)** Boxplot displaying estimated Neutrophil or naive B-cell frequencies in BL and w14 samples. In C the p-value is from a paired Wilcoxon rank-sum test; Samples of the same patient are connected with a grey line.

In responders, genes associated with inflammation, particularly neutrophil-driven (such as *DOK3, LRG1*, and *MMP9*), tended to be significantly less expressed in blood cells at w14 in comparison to BL, while upregulated genes were associated mostly with translation and other metabolic processes (eg. *EEF1A1*, RPL7, MRPL1, Fig. 4B). In agreement with this, serum proteins less abundant at w14 were associated with the activation of the complement system and innate immunity, including the complement factors CFB and CFH, and complement components C3, C8B and C8G (Fig. 4B, Supp. Table 2). Serum proteins more abundant at w14 were linked with vitamin metabolism, including the apolipoproteins APOA1, APOA2, and APOA4. Given the consistent decrease in expression of Neutrophil and innate immunity markers, we also compared estimated frequencies of different white blood cells between BL and w14. In agreement with the gene expression results, we observe in responders a significant decrease in neutrophil frequency at week 14, and an increase of B cell frequency (Fig. 4C), with a similar pattern observed for other adaptive immune cell populations such as CD4+ T-cells (Supp. Fig. 7A).

In non-responders, no significant differences in blood cell gene expression were found between BL and w14 (Supp. Fig. 3 and Supp. Table 3). Nonetheless, a rank-based gene set enrichment analysis (GSEA) of the transcriptomics data uncovers the same pathways as in responders (Supp. Fig. 4). In non-responders, 16 serum proteins were found differentially expressed (none with FC>2), of which 11 upregulated at w14, including APOA1, CLEC3B, CFH and RBP4 (Supp. Table 4). No significant pathways are found in the proteomics data. Also, although there is a similar tendency to decrease neutrophil frequency between BL and w14 in non-responders, it does not reach statistical significance in neutrophils nor other immune cell populations (Supp. Fig. 7A).

Genes and proteins differentially abundant between BL and w14 in responders were at more similar levels between the two patient groups at w14 (p<0.05), suggesting preexisting differences at baseline that got attenuated due to treatment (Supp. Fig. 5). In agreement with this observation, we did not find any genes or proteins displaying significantly different behavior between time and response group, suggesting that the action of adalimumab in responders and non-responders is similar. Overall, these results suggest that transcripts and proteins varying during adalimumab treatment were associated with a decrease in innate immune activity.

### Markers of inflammation were already lowered in the serum after 3-5 days of adalimumab treatment, in both responders and non-responders

To refine our understanding of the temporal response to adalimumab, we performed serum proteomics analysis also at 3-5 days and 2 weeks after beginning of treatment. In responders, among the serum proteins significantly downregulated at w14 in comparison to BL, Haptoglobin (HP), Haptoglobin receptor (HPR) and CRP showed a tendency to decrease already at 3-5 days, further decreasing until 2 weeks (Fig. 5A, Supp. Tables 5,6). These proteins also tended to decrease in non-responders, particularly in the first two weeks (Supp. Fig. 6A-C, Supp. Tables 7,8). Another group of proteins (including CFH and CFB) displayed a mild continuous decrease through time, while genes like C3 and C8A only appeared to decrease noticeably between week 2 and week 14. Among the proteins significantly increasing in w14 in comparison with BL in responders, there was greater heterogeneity, but some, including APOD, APOA2 and PPBP displayed a tendency to increase their abundance already at week 2, including in non-responders (Fig. 5B, Supp. Fig. 6D). Interestingly, the average level of change of these proteins was much milder (maximum FC of 2) when compared to HP, HPR and CRP (FC of 3-5). Thus, our results indicate that some markers of inflammation elevated at baseline were already lowered in the serum of some patients after just 3-5 days of adalimumab treatment.

**Figure 5:**
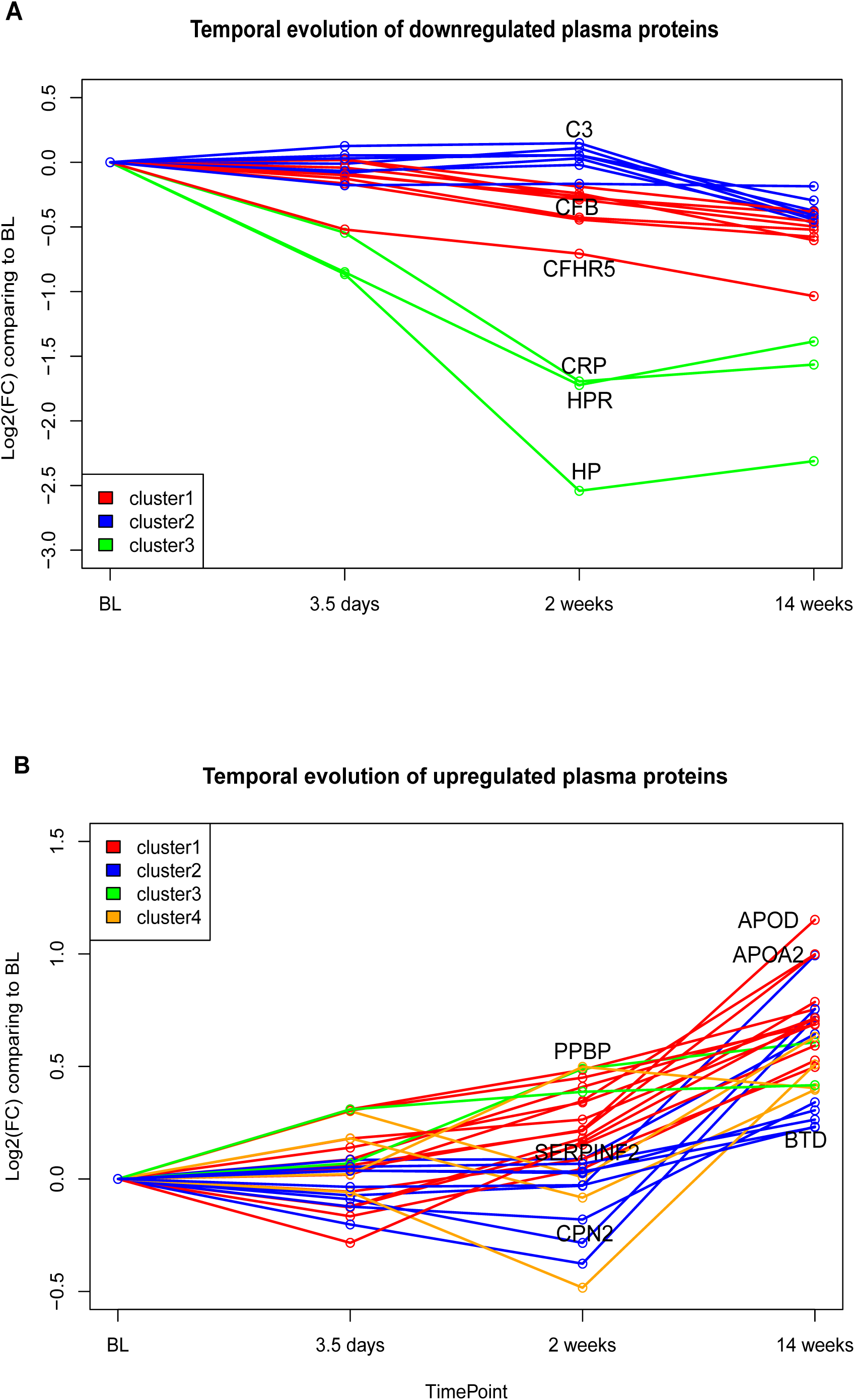
Markers of inflammation are already lowered in the serum after 3-5 days of adalimumab treatment, in both responders and non-responders. **A)** Log2 fold change of proteins between a given time point and the baseline. Only proteins significantly downregulated at w14 in responders were represented. **B)** Same as A but with upregulated proteins. We clustered proteins with similar temporal behavior using the dtwclust R package. Only the names of a set of representative proteins are displayed.

## DISCUSSION

This work documented that adalimumab treatment has a significant effect in transcript expression and protein abundances during the first 14 weeks of treatment. In our axSpA cohort, TNFi treatment seems to lower inflammatory markers in most patients, as observed in previous studies [4] and suggests an interplay between innate and adaptive immunity. Lymphoid markers such as *AFF3*, and Neutrophil/Lymphocyte ratios, emerge as novel differentiating variables between groups and enable highly accurate predictive models.

The main objective of our study was to identify molecular predictors, at BL, of response to TNFi. Our results corroborate ASDAS-CRP as an effective measure to decide, as promptly as possible, about TNFi as a therapeutic option in axSpA. However, the cut-off to predict response to therapy at baseline with maximum sensitivity is 4.1, representing a very high disease activity. Therefore, more variables are necessary for an accurate prediction, particularly in cases of moderate disease activity. Adding HLA-B27 status brought further improvements, while the addition of other variables (age at disease onset, disease duration) didn’t seem to add further resolution to the model. The observation of an interplay between innate and adaptive immunity, also reported in previous studies in RA [23], suggest similar mechanisms in both diseases. Our analysis suggests that the ratios between innate/adaptive immune populations, such as neutrophils/lymphocyte ratios deserve further exploration as a simple clinical marker with interest regarding TNFi therapeutic decision. In agreement with this, replacing HLA-B27 with the ratio of neutrophil frequencies over total lymphocytes at BL enabled an accurate model, although it didn’t seem to improve over HLA-B27. This suggests a potential clinical interest, particularly when HLA-B27 status is not easily available.

At the transcriptional level, our analysis revealed significant differences in several genes related to B cells (*AFF3, CD19, MS4A1, FCRLA, BANK1, PAX5*). In fact, the most enriched pathways were associated with B-cell development and activation (genes less expressed in responders), and to a lesser extent, neutrophil and inflammatory activity (eg. *RIPK3*, genes more expressed in responders). At the proteomic level, we could not detect any protein consistently different at baseline between responders and non-responders. One possible reason may be that B cell proteins are present at too low a level in serum, which combined with the overall heterogeneity of the samples, made these undetectable in our proteomic analysis. We then used RNA-Seq data to estimate frequencies of different white blood cell populations that are not provided in the normal clinical hemogram, such as B-Cells. Our analysis confirmed a higher frequency of neutrophils, and a lower relative frequency of B cells (and of other adaptive immune populations) in responders in comparison with non-responders at baseline. Adding *AFF3*, a B-cell associated gene that was the top differentially expressed gene at BL between responders and non-responders, in a logistic regression model with ASDAS-CRP, enabled a very accurate prediction of response to adalimumab (AUC=0.96). A similarly high predictive capacity was obtained with another B-cell associated gene (*BANK1*) providing further consistency to our results. Moreover, using the same models with robust machine learning methods including cross-validation suggests a predictive capacity of over 80% accuracy, an estimate which is likely to be very conservative given the small size of our cohort.

To provide further insights regarding the molecular mechanisms involved in the differential response to TNFi treatment, we obtained measures of expression for blood transcripts and serum proteins of axSpA patients during 14 weeks of treatment with adalimumab. Our transcriptomics and proteomics data indicate that a significant fraction of the observed variation in gene expression can be explained by treatment time and response status. Moreover, transcripts and proteins with significantly reduced expression between BL and w14 of treatment were associated with inflammation and innate immunity, in agreement with the observed changes in clinical markers of inflammation and scores of disease activity. Taken together, this suggests the existence of clinically relevant information in the data and the potential to uncover early biomarkers of TNFi response.

Our transcriptomic results are in large agreement with previous results in axSpA. Namely, *TNFSF14* (LIGHT), *IL17RA*, and *EPOR*, genes reported by Haroon et al. [16], also had significantly reduced expression after TNFi treatment in our cohort (the gene *IFNAR1* didn’t reach significance in our study but had a borderline adjusted p-value of 0.07). Moreover, 58 (16%) of 360 genes upregulated after TNFi in Wang et al. [17] were also upregulated in our study, and 88 (31%) of 285 down regulated genes were also detected in our study (with only one gene showing opposing significant tendencies between the studies). Among the 103 significantly differentially expressed genes (DEGs) between BL and w14 with higher fold changes (FC>2), ten had been previously identified as axSpA-associated in GWAS [35]. Of these, *TNFRSF1A, TBKBP1, HHAT*, and *LTBR*, all less expressed at w14, are involved in the TNF pathway, mediate apoptosis through nuclear factor-κB, and function as a regulator of inflammation [36]. Interestingly, *IL1R, IL6R* and *TYK2*, more associated with innate immunity, were downregulated, while *IL7R* and *ICOSLG*, more associated with the adaptive immune system, namely the stimulation and differentiation of T and B cells, were upregulated. *FCGR2A*, also downregulated, encodes a cell surface receptor found on phagocytic cells, such as, macrophages and neutrophils. This indicates that, despite heterogeneity in clinical manifestations of axSpA, molecular response to TNFi seems consistent between different studies, at least at the level of the blood cell transcriptome. These results suggest that biomarkers of TNFi response uncovered in our cohort are likely to be generally applicable in the axSpA context, although further validation is still mandatory.

Our study expands on previous works in the axSpA context by introducing proteomics data, including intermediate timepoints during adalimumab treatment. We identified several serum proteins undergoing significant changes in abundance with adalimumab treatment. Overall, we observe a decrease in the abundances of HP, HPR, CRP and complement factors, and an increase of several apolipoproteins, CLEC3b and RBP4. Interestingly, in responders, an early (since days 3-5) and persistent decrease of HP and CRP is seen, correlating with observed clinical improvement. Decrease of complement factors is milder and occurs later (after week 2). A later increase of apolipoproteins APOA1, APOA2 and APOD, involved in lipid clearance from circulation and anti-inflammatory properties, has been previously shown [37]. Curiously, APOD has no marked similarity to other apolipoprotein sequences, but has a high degree of homology to plasma retinol-binding protein (Rbp), which is also overexpressed, and both are thought to influence bone metabolism [37,38]. Rbp4 seems to be present in a restricted population of epiphyseal chondrocytes and perichondrial cells correlating to the future regions of secondary ossification. In addition, CLEC3B, a plasminogen-binding protein induced during the mineralization phase of osteogenesis, is also more abundant [39]. These data unravel molecular mechanisms underlying the abrogation of inflammation, while also suggesting that osteoproliferation may be induced under TNFi therapy, as documented in recent studies [40–42]. Future studies should aim to uncover the physiopathological role of these genes in TNFi response.

We acknowledge limitations in our study, namely the need for further validation with targeted approaches and in new cohorts. Moreover, as all patients were under adalimumab treatment, we cannot extrapolate the results to other TNFi, although comparison with analysis in RA suggest that the underlying mechanisms will be similar.

## CONCLUSIONS

To our knowledge, this study is the first using a multi-omic approach to tackle the difficult challenge of predicting at BL the therapeutic response to TNFi in the axSpA context. Our results suggest an interplay between innate and adaptive immunity occurring under TNFi therapy, with lymphoid markers emerging as the most differentially expressed between groups and enabling highly accurate predictive models with our cohort. In addition, our work confirms transcriptomics results of previous studies investigating the effects of TNFi in axSpA and expands them by providing a genome-wide census of both blood cell genes expression and serum proteins abundances during the first 14 weeks of treatment with Adalimumab, in both responders and non-responders.

Taken together, our results suggest that molecular data can not only provide mechanistic insights to the genesis and progression of the disease, but also suggest novel biomarkers to evaluate the potential response to adalimumab before initiating treatment or in its initial phases.

## Supporting information

Sup material description

SuppFig1

SuppFig2

SuppFig3

SuppFig4

SuppFig5

SuppFig6

SuppFig7

SuppFig8

SuppFig9

SuppTable1

SuppTable2

SuppTable3

SuppTable4

SuppTable5

SuppTable6

SuppTable7

SuppTable8

SuppTable9

SuppTable10

SuppTable11

SuppTable12

## Data Availability

Transcriptomics and proteomics data are available as supplementary tables in zenodo under the DOI https://doi.org/10.5281/zenodo.4914646. Mass spectrometry data are available through the ProteomeXchange Consortium via the PRIDE partner repository (https://www.ebi.ac.uk/pride/), under dataset identifiers PXD026189 and http://doi.org/10.6019/PXD026189.

## DECLARATIONS

### Ethics approval and consent to participate

This study (registered in clinical trials.gov with the identifier NCT02492217) was approved by the national competent authorities: National Ethics Committee for Clinical Research (CEIC) and INFARMED (competent authority to regulate medicinal products). The study was conducted according to Good Clinical Practices (GCP), Declaration of Helsinki, and legal regulation applicable, ensuring participants’ safety and mechanisms for data protection. Written informed consent was obtained from all participants before study inclusion. None of the patient identifiers were known to anyone outside of the research group. There was no active involvement of patients or the public as co-producers of research in this project. However, all participants were invited to participate in a meeting where the main results were presented and discussed.

### Author’s Contributions

Study concept and design: F.M.P.-S., J.C.B.; Acquisition of clinical data: M.B., P.P., H.S., J.L.-G., J.T.-C., J.A.P.S., J.M.D., A.B., S.M., J.C.B., F.M.P.-S.; Laboratorial research direction: A.V.C., F.M.P.-S.; Performance of laboratory experiments: A.S., A.F.F., F.M.P.-S., A.V.C., J.-C.G., J.A., V.B.; Analysis and interpretation of data: D.S., A.V.C., A.F.F., F.M.P.-S.; Writing of the manuscript: D.S., A.V.C., A.F.F., F.M.P.-S.; Critical revision of the manuscript for important intellectual content: All authors.

## Acknowledgments

We thank the members of the computational multi-omics group for critical reading of the document. We acknowledge the careful comments provided by 2 anonymous reviewers in a previous submission.

## Abbreviations

ASAS: assessment in ankylosing spondylitis
ASDAS: ankylosing spondylitis disease activity score
AUC: area under the curve
axSpA: axial spondyloarthritis
BASDAI: bath ankylosing spondylitis disease activity index
bDMARD: biological disease-modifying antirheumatic drugs
BL: baseline
ESR: erythrocyte sedimentation rate
FC: fold change
GSEA: gene set enrichment analysis
HLA-B27: human leukocyte antigen B27
MRI: magnetic resonance imaging
NSAID: non-steroidal anti-inflammatory drugs
PCA: principal component analysis
RA: rheumatoid arthritis
sPLS-DA: sparse partial least squares discriminant analysis
TNFi: tumor necrosis factor inhibitor
w14: week 14

