## Supplementary material for "MULTI-OMICS TEMPORAL PROFILING OF AXIAL SPONDYLOARTHRITIS PATIENTS REVEALS AN ASSOCIATION OF THERAPEUTIC RESPONSE TO ADALIMUMAB WITH DISEASE ACTIVITY AND INNATE / ADAPTIVE IMMUNITY": Sup material description

**Supplemental Material**

**Criteria For Patients selection**

Inclusion Criteria:

●	axSpA according to Portuguese Society of Rheumatology (SPR) guidelines (1984 modified New York Criteria, but allowing the use of MRI as imaging criteria)

●	Patient enrolment followed national guidelines for TNF antagonist use for the treatment of axSpA

●	Adults between 18 to 75 years

●	Ability to provide informed consent

●	Corticosteroid therapy allowed (equivalent to ≤ 10 mg prednisone) and / or NSAID (nonsteroidal anti-inflammatory drug), stable dose in 4 weeks before study initiation

●	Adequate contraception (barrier or hormonal) in men and women of childbearing age (patients and their partners)

●	Adequate renal and hepatic function (2 times ULN)

Exclusion Criteria:

●	Current pregnancy or breastfeeding

●	Previous treatment with biologic DMARD's (disease-modifying antirheumatic drug)

●	Intraarticular (including sacroiliac joints) and periarticular injections within 28 days before screening.

●	History of rheumatic disorder other than axSpA

●	Other forms of spondylarthritis than axSpA

●	Any uncontrolled medical condition (e.g., uncontrolled diabetes mellitus, unstable ischemic heart disease)

●	History or signs of demyelinating disease

●	Malignancy (except for completely treated squamous or basal cell carcinoma)

●	Moderate to severe heart failure (NYHA class III/IV) Positive serology for hepatitis B, hepatitis C, or human immunodeficiency virus

●	Active or latent tuberculosis (TB) or histoplasmosis or other severe infections such as sepsis, and opportunistic infections

●	Infections requiring hospitalization or intravenous treatment with antibiotics within 30 days or oral treatment with antibiotics within 14 days before enrollment

●	Ankylosis of the spine (syndesmophytes presence at all levels from D12 to S1 in X-ray (XR) lateral view)

●	Hypersensitivity to the active substance or to any of the excipients

**Supplemental Figure Legends**

**Supplemental Figure 1: The TNF inhibitor adalimumab induces a reduction in clinical disease parameters. A)** Paired values of ASDAS-CRP scores, separated by response groups. Samples from the same patient are connected with a grey line between timepoints. **B)** Paired values of BASDAI scores, separated by response groups. In all cases, the differences were statistically significant (p<0.05), as estimated using a paired Wilcoxon signed rank test.

**Supplemental Figure 2: Response to TNFi treatment is a significant factor distinguishing responders and non-responders at baseline.** Sparse partial least squares discriminant analysis (sPLS-DA) of proteomics data in responders (AUC=1, p=1.1e-05) and non-responders (AUC=1, p=3.4e-05) using time as a variable of interest; In all cases, AUC and p-value correspond to the two best components of the sPLS-DA. In all graphs, ellipses represent 95% confidence intervals.

**Supplemental Figure 3: Non-responders have fewer robustly differentially expressed genes/proteins between BL and W14.** Volcano plot (log2 of the fold change versus -log10 of the unadjusted p-value) comparing the transcriptomics (Supp. Table 3) and proteomics (Supp. Table 4) baseline samples versus week 14 samples in non-responders; for visual purposes, the p-values in the transcriptomics plot were plotted, and not the adjusted p-values as these were all 1 or very close to 1; non-significant (NS) genes/proteins are in grey; in blue genes/proteins that are statistically significant but have a mild fold change (less than 2); some of the blue genes names are displayed in the plot.

**Supplemental Figure 4: TNFi treatment in non-responders acts in the same pathways as responders.** Barplot displaying the Normalized Enrichment Score (NES) of representative significant pathways resulting from a gene set enrichment analysis (GSEA) comparing transcript expression of week 14 (W14) against baseline (BL) non-responder samples.

**Supplemental Figure 5: Transcriptomic and proteomic differences detected between BL and W14 in responders are attenuated between responders and non-responders at W14.** Box plot of the log2 fold change of genes: **A)** “R: W14 vs BL” are w14 samples against baseline samples in responders; “W14: R vs NR” are responder versus non-responders at w14, Only the genes differentially expressed from “R: W14 vs BL” are represented. **B)** Same as A but regarding non-responders. **C)** Same as A but regarding proteins. **D)** Same as A but regarding proteins in non-responders. In all cases p<0.05.

**Supplemental Figure 6: Markers of inflammation are already lowered in the serum after 3-5 days of adalimumab treatment, in both responders and non-responders. A)** Mean clinical CRP values in different time points, in responders and non-responders; Mean spectral counts per million (sCPM) values in different time points, in responders and non-responders in **B)** CRP; **C)** HP; **D)** APOA2. Error bars represent standard deviation.

**Supplemental Figure 7: Blood transcriptome data at baseline suggests that response to adalimumab derives from an interplay between innate and adaptive immunity. A)** Heatmap representation of fold change values from a quantitative set analysis for gene expression (QUSAGE) used with immune signature gene sets from Lewis et al. W14.R_BL.R indicates comparison of week 14 samples against baseline samples in responders; W14.NR_BL.NR indicates comparison of week 14 samples against baseline samples in non-responders; BL.R_BL.NR indicates comparison of responders against non-responders at baseline; W14.R_W14.NR indicates comparison of responders against non-responders at week 14 (* indicates p-value of fold-change < 0.05; ** p-value < 0.01; *** p-value < 0.001); **B)** Heatmap representation of a cross-correlation analysis of the relative frequencies of different immune populations and clinical parameters; only significant correlations are displayed. Highly correlated variables are clustered together.

**Supplemental Figure 8: RNA-Seq derived estimates of white blood cell populations correlate well with values from clinical hemograms. A)** Correlation between Lymphocyte frequencies derived from RNA-Seq or from clinical hemograms (Pearson R=0.86, p=1.16e-8, n=27) **B)** Correlation between Neutrophil frequencies derived from RNA-Seq or from clinical hemograms (Pearson R=0.84, p=3.36e-8, n=27) **C)** Ratio between Neutrophil and Lymphocytes derived from hemograms at baseline (Wilcoxon rank-sum test p=0.006, responders n=10, non-responders n=8) **D)** Ratio between Neutrophil and Lymphocytes derived from hemograms at week 14 (Wilcoxon rank-sum test p=0.79, responders n=7, non-responders n=4). **E)** Ratio between Neutrophil and Lymphocytes derived from Cibersort estimates at baseline (Wilcoxon rank-sum test p=0.008, responders n=16, non-responders n=17). **F)** Ratio between Neutrophil and Lymphocytes derived from Cibersort estimates at week 14 (Wilcoxon rank-sum test p=0.78, responders n=18, non-responders n=17). Colors represent response group and time point.

**Supplemental Figure 9: Clinical CRP measures correlate well with CRP expression measurements from the proteomics assay.** Correlation between clinical CRP and the spectral counts per million (sCPM) values for CRP in the proteomics data. Spearman rho = 0.79; Pearson correlation on the log scale = 0.72; in both cases p-value<e-16. Values were log transformed, as the correlation seemed non-linear (the spearman correlation is the same as in the linear scale, and pearson correlation is still 0.64, p=2.9e-13).

**Supplementary Table 1**. **Results of the differential expression analysis comparing the transcriptomics of responders between week 14 and baseline.** For each gene we indicate the estimated log2(Fold Change), average expression (across all samples), moderated t-statistic, p-value, adjusted p-value, and B-statistic (posterior log-odds of differential expression). Genes are presented ordered by the adjusted p-value.

**Supplementary Table 2.** **Results of the differential expression analysis comparing the proteomics of responders between week 14 and baseline.** Same as Supplementary Table 1, but for proteomics data.

**Supplementary Table 3.** **Results of the differential expression analysis comparing the transcriptomics of non-responders between week 14 and baseline.** Same as Supplementary Table 1, but for non-responders.

**Supplementary Table 4.** **Results of the differential expression analysis comparing the proteomics of non-responders between week 14 and baseline.** Same as Supplementary Table 1, but for proteomics data in non-responders.

**Supplementary Table 5.** **Results of the differential expression analysis comparing the proteomics of responders between 3-5 days and baseline.** Same as Supplementary Table 1, but for proteomics data, comparing time-point 2 (3-5 days) with baseline, in responders.

**Supplementary Table 6.** **Results of the differential expression analysis comparing the proteomics of responders between 2 weeks and baseline.** Same as Supplementary Table 1, but for proteomics data, comparing time-point 3 (2 weeks) with baseline, in responders.

**Supplementary Table 7.** **Results of the differential expression analysis comparing the proteomics of non-responders between 3-5 days and baseline.** Same as Supplementary Table 1, but for proteomics data, comparing time-point 2 (3-5 days) with baseline, in non-responders.

**Supplementary Table 8.** **Results of the differential expression analysis comparing the proteomics of non-responders between 2 weeks and baseline.** Same as Supplementary Table 1, but for proteomics data, comparing time-point 3 (2 weeks) with baseline, in non-responders.

**Supplementary Table 9.** **Results of the differential expression analysis comparing the proteomics at baseline between responders and non-responders.** Same as Supplementary Table 1, but for proteomics data, comparing responders with non-responders at baseline.

**Supplementary Table 10.** **Results of the differential expression analysis comparing the transcriptomics at baseline between responders and non-responders.** Same as Supplementary Table 1, but comparing responders with non-responders at baseline.

**Supplementary Table 11. RNA-Seq gene counts from kallisto.**

**Supplementary Table 12. List of proteins identified by proteomics and their abundances estimated by spectral counts.**
