## Supplementary figures and images for "MULTI-OMICS TEMPORAL PROFILING OF AXIAL SPONDYLOARTHRITIS PATIENTS REVEALS AN ASSOCIATION OF THERAPEUTIC RESPONSE TO ADALIMUMAB WITH DISEASE ACTIVITY AND INNATE / ADAPTIVE IMMUNITY"

### SuppFig1

**Supplemental Figure 1**

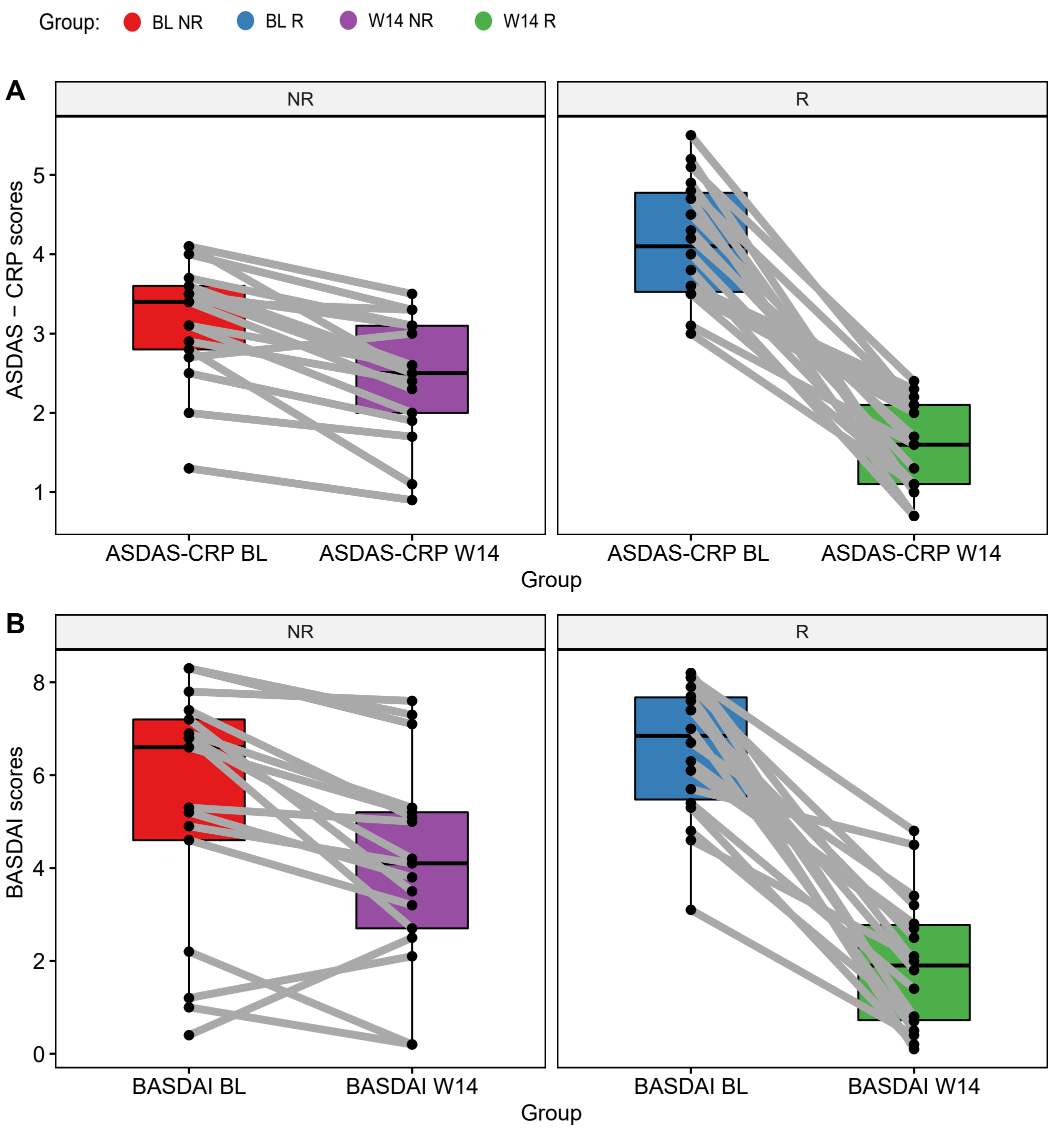

### SuppFig2

Supplemental Figure 2

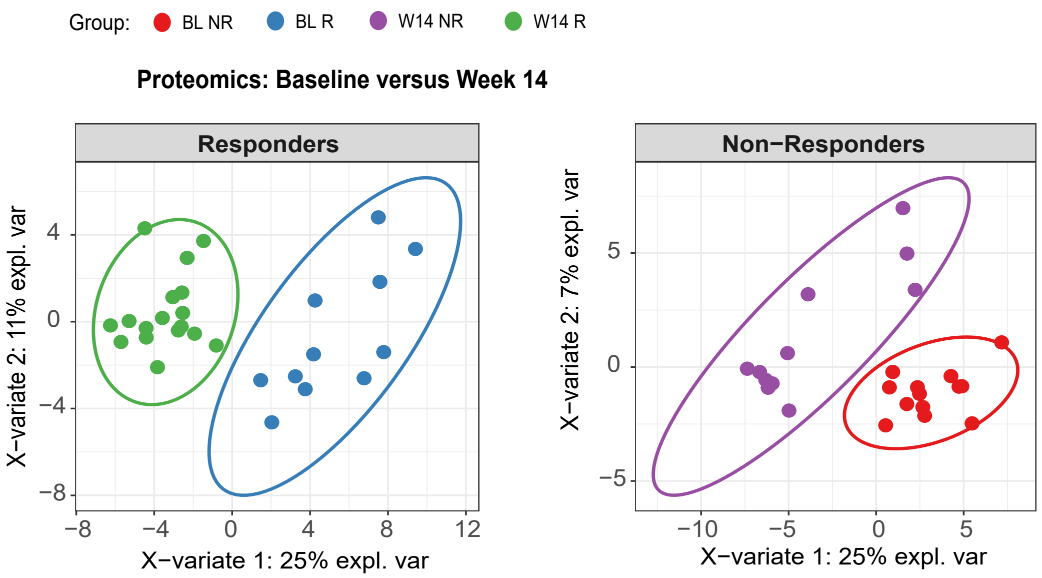

### SuppFig3

**Supplemental Figure 3**

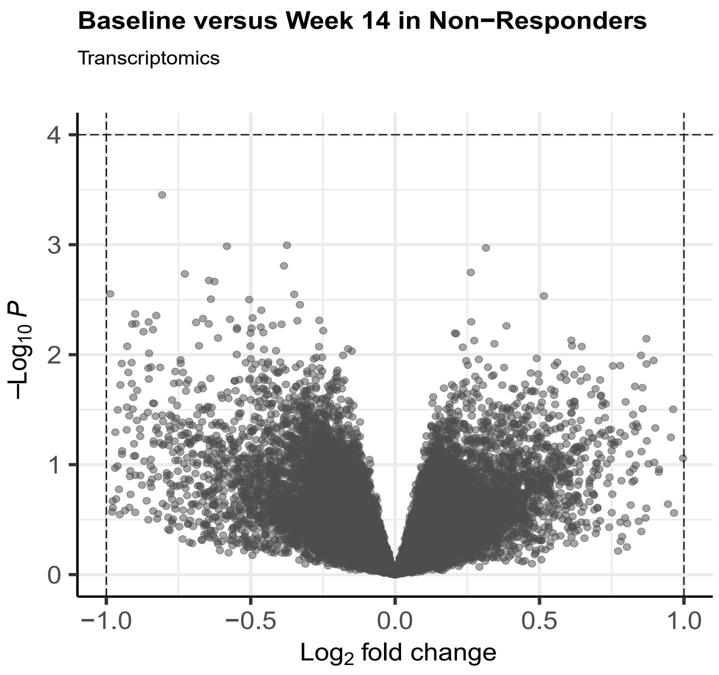


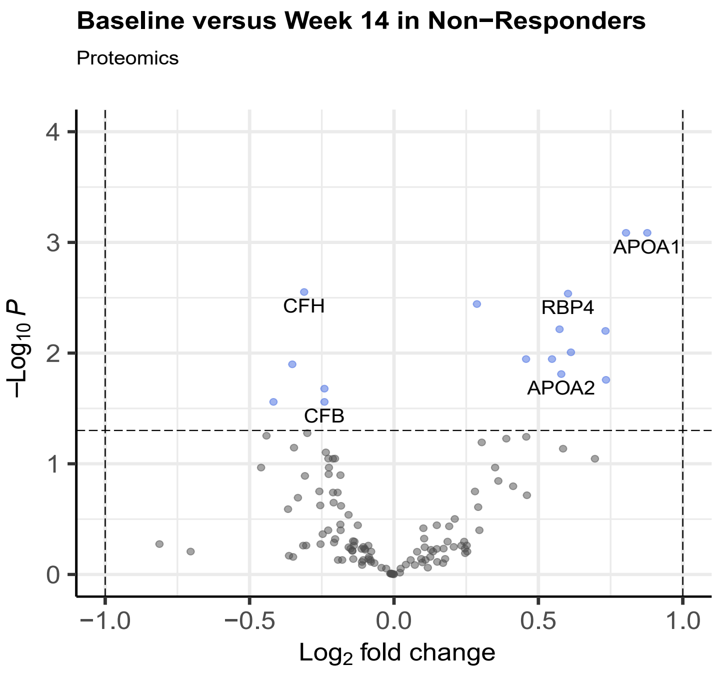

### SuppFig4

**Supplemental Figure 4**

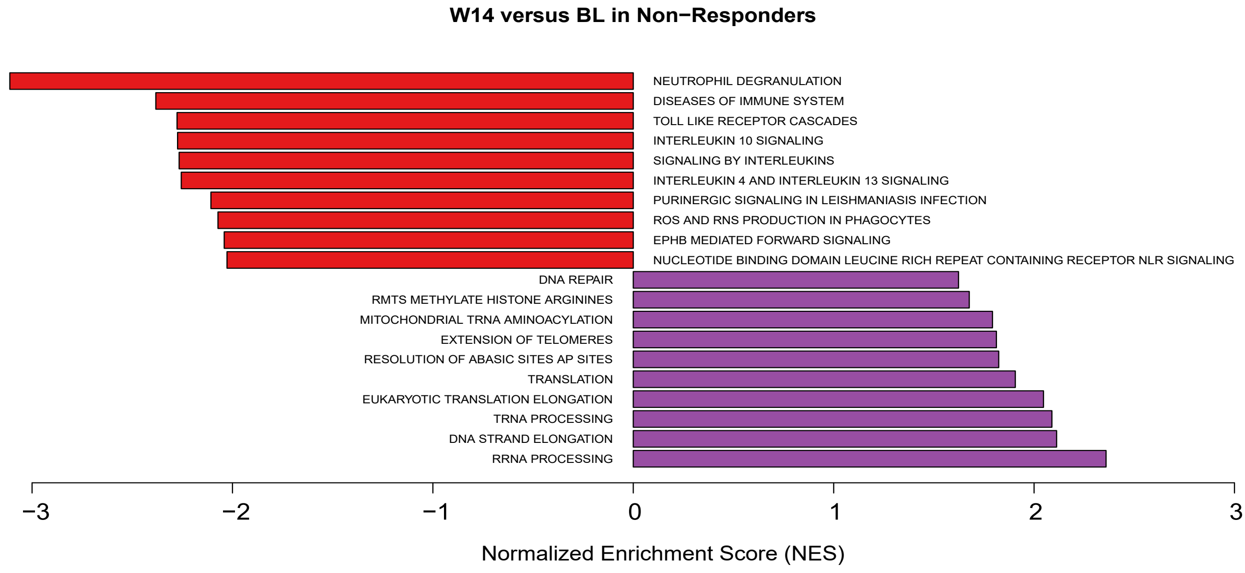

### SuppFig5

Supplemental Figure 5

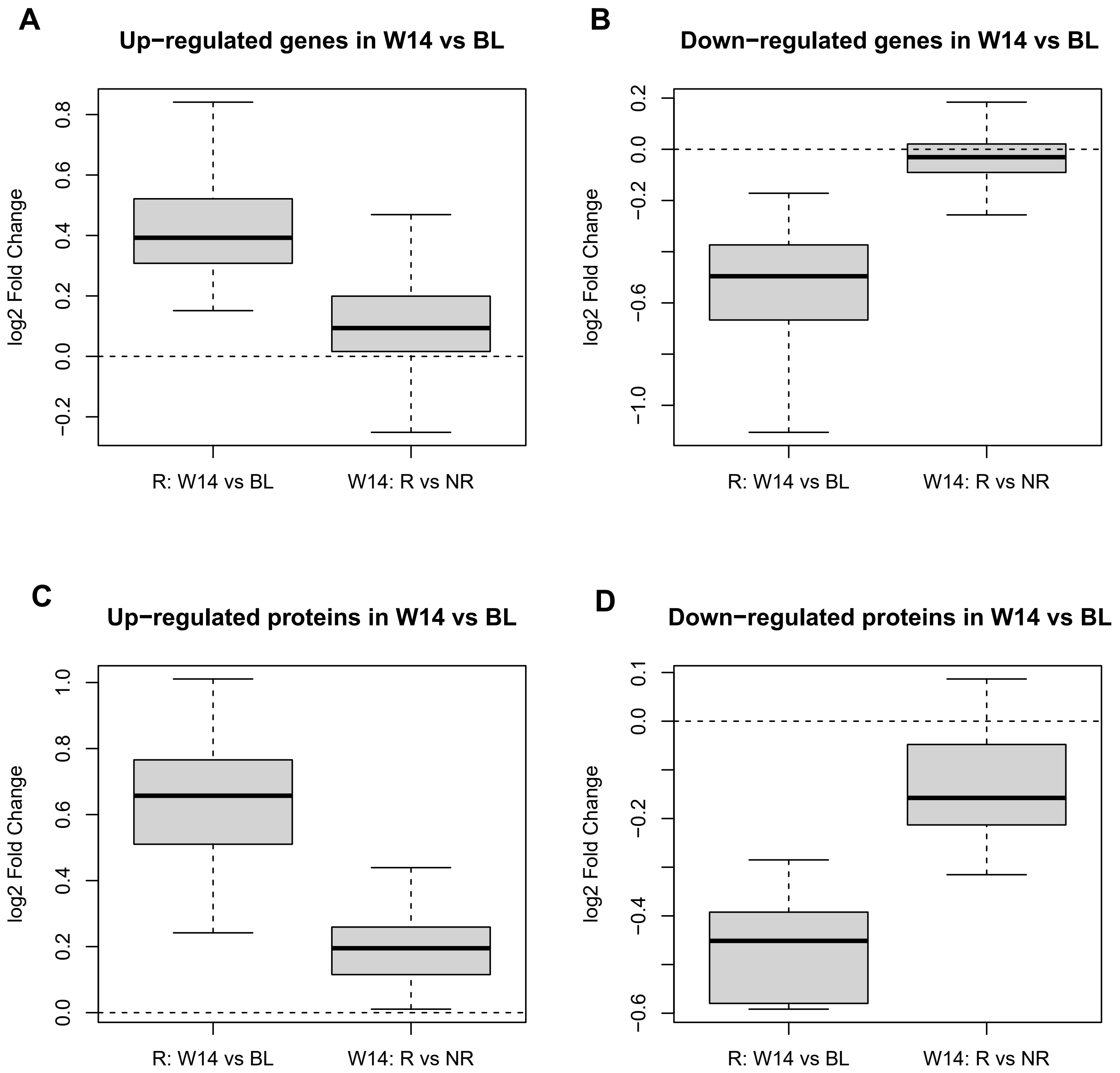

### SuppFig6

**Supplemental Figure 6**

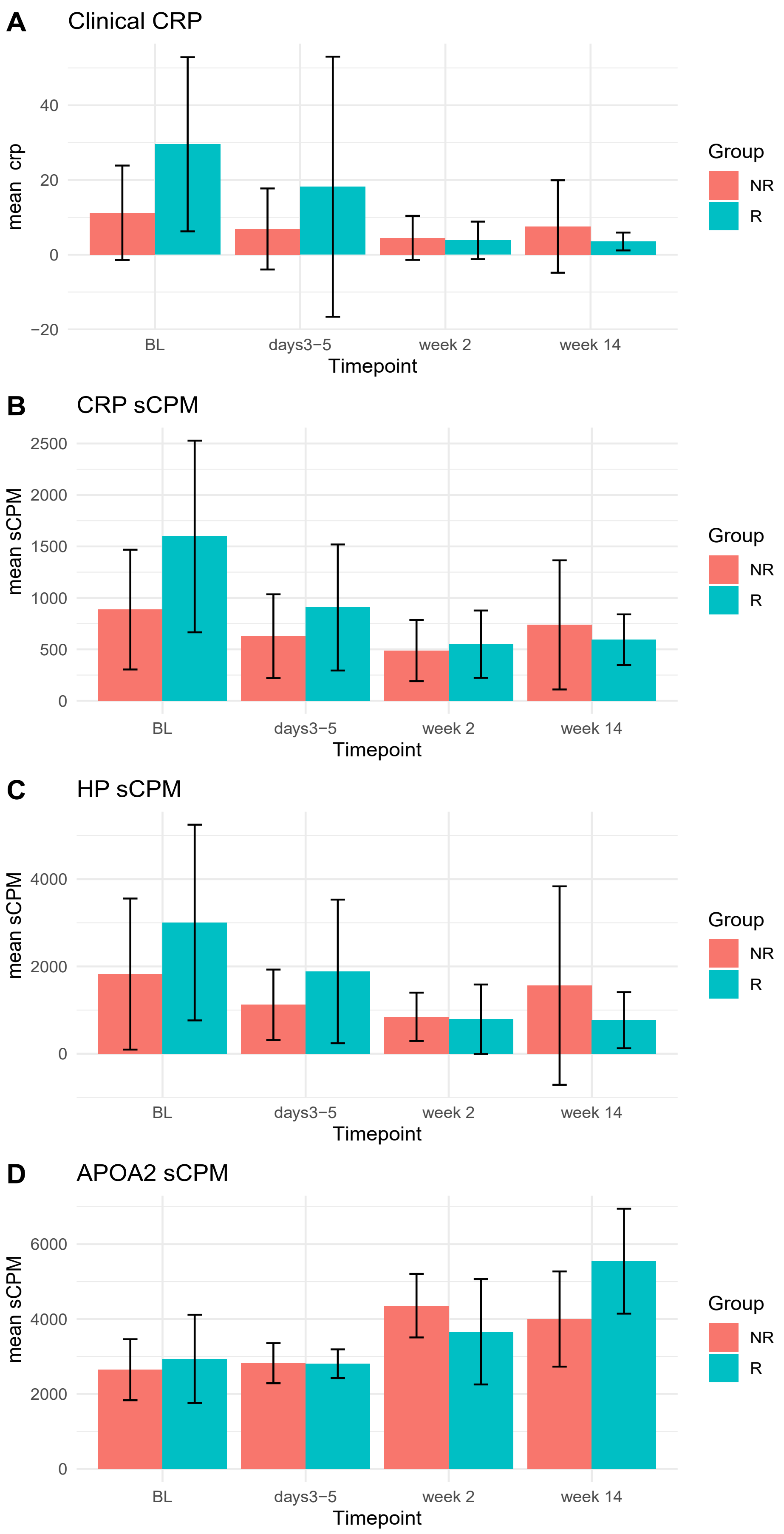

### SuppFig7

**Supplemental Figure 7**

A

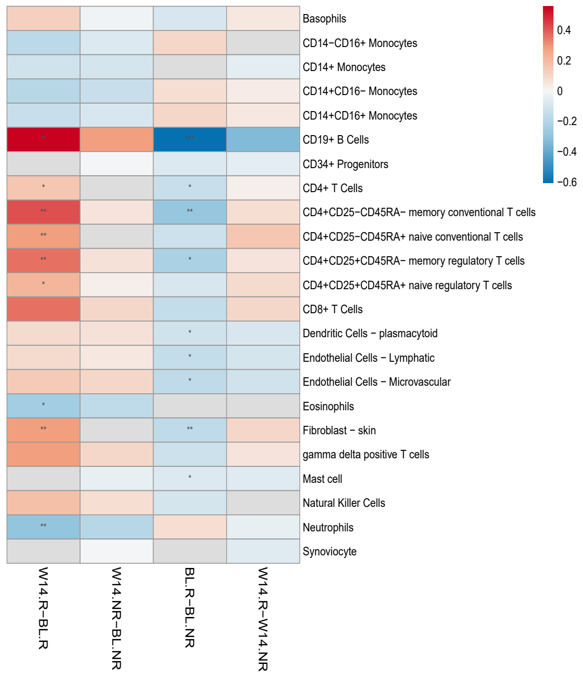


B

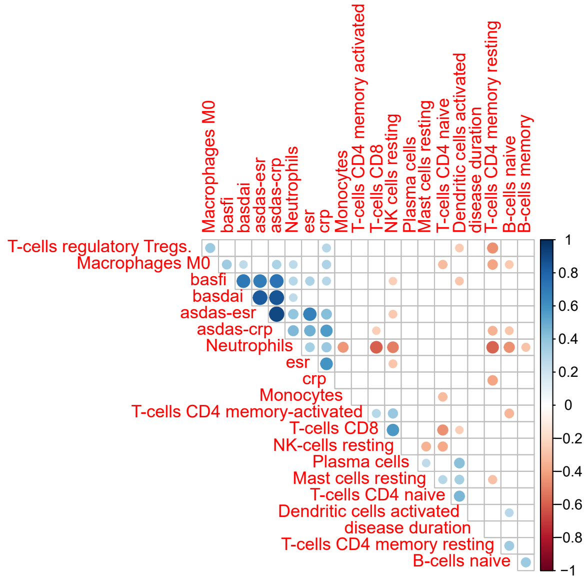

### SuppFig8

Supplemental Figure 8

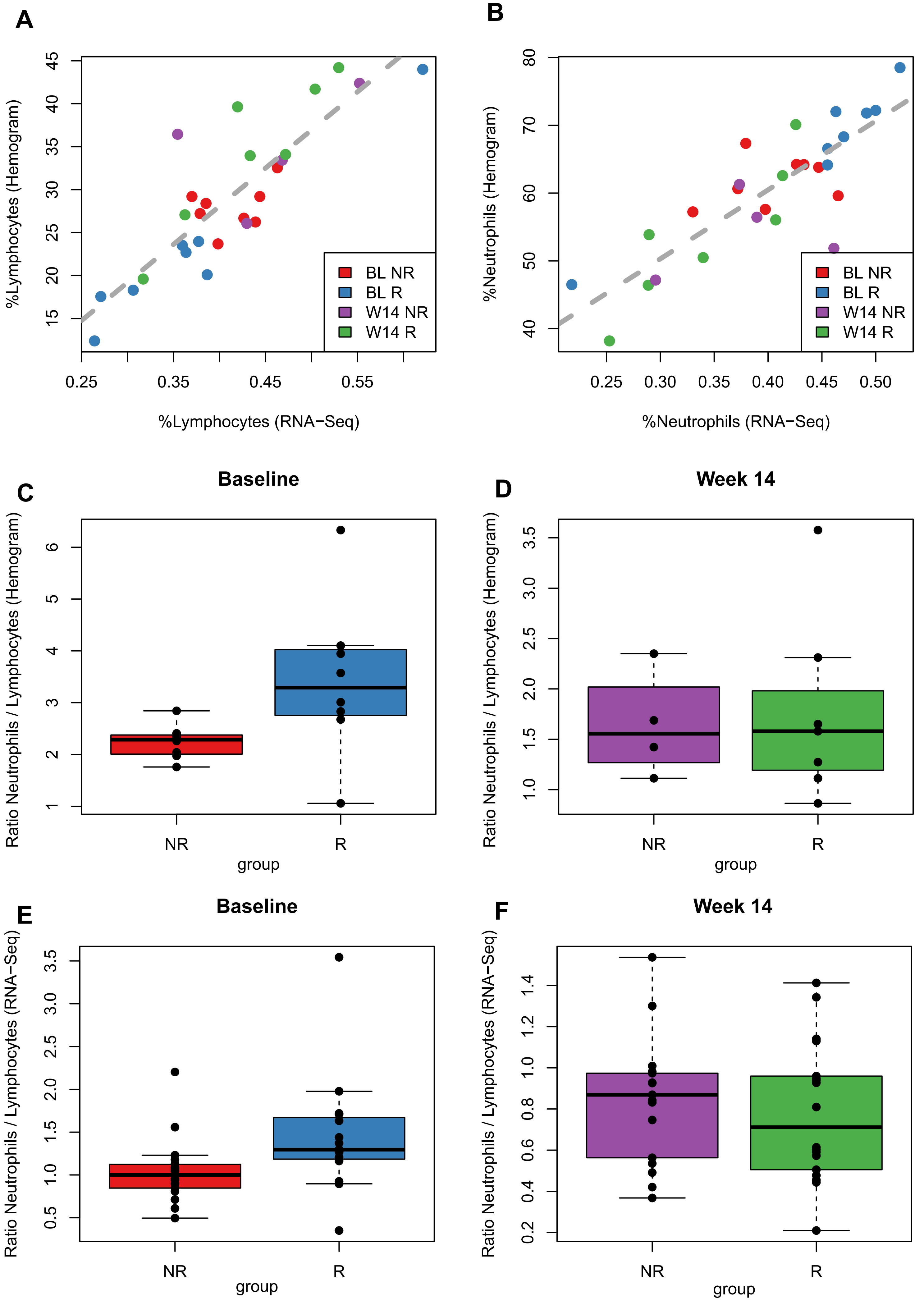

### SuppFig9

**Supplemental Figure 9**

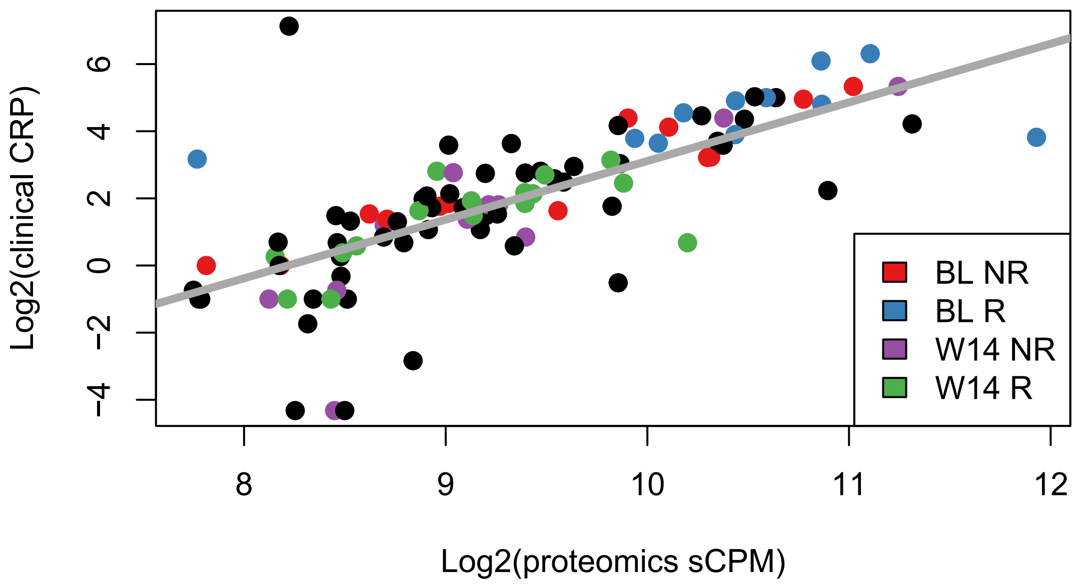
